# Dietary Enhancement for Iron Deficiency and Anaemia Treatment (DEFeAT): study protocol for a randomised controlled trial to assess the effect of a mineral-enriched powder on iron deficiency in women of reproductive age

**DOI:** 10.64898/2026.10.20.26365734

**Authors:** Jessie L Burns, Hailey Scott, Lawrence Mbuagbaw, Clara H Miller, Hauna Sheyholislami, Brianne A Clouthier, Bénédicte Fontaine-Bisson, Kristin L Connor

## Abstract

**Background:** Iron deficiency (ID) and iron deficiency anaemia (IDA) have global and severe health implications, and disproportionately impact women of reproductive age. Current methods of supplementing diets with iron are challenged by poor adherence and palatability, reducing effectiveness of these treatment strategies. The aim of this study is to evaluate the efficacy of a novel home beverage fortification with a mineral-enriched powder, designed to enhance bioavailability and ease of use, for the treatment of ID in women of reproductive age.

**Methods:** This single-centre, randomised, double-blinded placebo-controlled trial will enrol 150 eligible biologically female participants with ID. Participants will be randomly assigned to the active intervention (iron/zinc-enriched powder) or control group (placebo powder) over 6 months. Primary outcomes will include change in serum ferritin (SF) concentrations and proportion of participants who remain iron deficient at 6-months. Secondary outcomes will include change in SF and haemoglobin (Hb) concentrations across timepoints, and levels of metabolites and key circulating pro-inflammatory biomarkers, changes in dietary intakes, self-reported perceptions of health, barriers to use of the powder, adherence and palatability, safety outcomes, and cost-effectiveness. Exploratory outcomes will assess the gut microbial metagenome and metabolome.

**Discussion:** Fortification, an effective approach for the prevention of ID/IDA, has advantages over some methods of supplementation because it uses low levels of iron, minimising unpleasant side effects, does not risk toxicity, and has high acceptability and compliance rates. This mineral-enriched powder uses electrolytic iron and zinc sulphate monohydrate of a small particle size, which is less irritating to the gut, and facilitates bioavailability. Findings from the study will be shared with those affected by and involved in treating these conditions, as a novel intervention that is both effective and acceptable among women of reproductive age would help to improve treatment compliance, and ultimately, reduce ID/IDA burden.

**Trial registration:** This trial is registered at clinicaltrials.gov (NCT05990166): https://clinicaltrials.gov/study/NCT05990166, August 2023.

## Introduction

### Background and rationale [6a]

Micronutrient deficiencies, particularly iron deficiency (ID), are a serious global health concern associated with various adverse health issues^1^. ID globally affects 20-25% of the population, and 1 in 3 women worldwide^2^. While micronutrient deficiencies are particularly prevalent in low- to middle-income countries (LMICs) due to the high burden of hidden hunger in these regions^3,4^, ID continues to persist in developed countries despite the existence of available treatment options^5,6^. If left untreated, ID can lead to iron deficiency anaemia (IDA), a condition characterised by chronically low iron levels in the body. ID is recognised as the leading cause of anaemia globally according to the World Health Organization (WHO), with the global prevalence of anaemia showing only a marginal decrease over the past three decades across both developed and LMICs (from 28-24% from 1990 to 2021)^6^. Failure to address ID/IDA can result in severe long-term adverse health outcomes in affected individuals, including weakened immune function, impaired cognitive ability, and a higher susceptibility to morbidity from infections^7,8^.

Women of reproductive age (18-49 years) and pregnant people are at highest risk for developing ID and IDA due to their increased biological demand for iron to support menstruation and pregnancy^9–11^. In the non-pregnant state, adequate amounts of iron are needed to support disease resistance, cognitive function, and reproductive health^12–14^. During pregnancy, individuals have an increased physiological need for iron to support both their own health and fetal development^7,15,16^. Globally, 37% of women and 30% of pregnant people are affected by ID and IDA^17–20^. For pregnant people, ID and IDA can be particularly dangerous, as it is associated with an elevated risk of maternal death, preterm birth, and low birthweight in offspring^7,21–23^. Given the substantial prevalence and adverse consequences of untreated ID for women of reproductive age and pregnant people, there is an urgent need for novel, sustainable, and effective treatment strategies to combat this public health issue.

Zinc is another nutrient that is important for health as it is involved in numerous enzymatic processes, including those regulating iron metabolism^24–26^. Consequently, zinc deficiency may contribute to the development or exacerbation of ID^24–26^. ID and zinc deficiency often occur together, as they are found in many of the same foods and share similar absorption pathways in the small intestine; thus, zinc deficiency should also be considered in the management of ID^24,27^.

The standard of care treatment for ID involves oral iron therapy with supplemental iron^11,28^. However, compliance rates with iron supplementation are low (often less than 50%)^29^, which ultimately reduces the overall effectiveness of treatment. Alternatively, ID can be treated with fortification of food^30^. Studies have demonstrated the effectiveness of food fortification in treating ID and IDA by improving clinical outcomes, including increased serum ferritin (SF) and haemoglobin (Hb) concentrations^31–35^. Although fortification is less immediate in alleviating these deficiencies, it presents several advantages over supplementation, including fewer side-effects, no risk of toxicity, and high acceptability and compliance rates among end-users^34,36,37^.

In recent years, innovative fortification products have emerged as promising treatments for ID and IDA^38–40^. These products include micronutrient powders (MNPs; such as ‘Sprinkles’^41,42^) and iron ingots^35^, allowing for at-home iron fortification. However, iron compounds are known to pose challenges for fortifying foods given their tendency to react strongly with many food constituents, leading to poor palatability and thus low long-term adherence^36,43,44^. This could likely be one of several factors explaining overall, relatively low adequate adherence rates to MNPs^45,46^. Thus, there is a need to develop and implement palatable, effective, and acceptable MNP interventions tailored for women of reproductive age^11,47^.

To fill this need, a novel iron- and zinc-enriched powder was developed to allow for at-home fortification. This mineral-enriched powder is a proprietary mixture of electrolytic iron and zinc sulphate monohydrate of a small particle size, intended to enhance bioavailability. This powder can be conveniently added directly to beverages, overcoming various barriers associated with traditional food fortification methods. As a first step, we will test the efficacy of the mineral-enriched powder in reproductive aged women in a high-income country where ID is prevalent. Future trials will consider groups in LMICs with high prevalence of both iron and zinc deficiencies.

## Objectives [7]

### 1. Primary objectives

To compare the effect of six months of home beverage fortification with an iron and zinc enriched powder versus placebo in iron deficient reproductive aged women (serum ferritin [SF] ≥12 µg/L and ≤30 µg/L) on:

1. Change in SF concentrations at six months (endpoint) (primary clinical outcome)
2. Proportion of participants who are no longer iron deficient at six months (primary clinical outcome)

### 2. Secondary objectives

To compare the effect of six months of home beverage fortification with an iron and zinc enriched powder versus placebo in iron deficient reproductive aged women on:

#### a. Clinical outcomes

i. Change in SF and haemoglobin (Hb) concentrations from baseline, to midline (three months) and endline (six months)
ii. Levels of metabolites (circulating metabolome) at baseline, three months, and six months
iii. Levels of key circulating pro-inflammatory biomarkers at baseline, three months, and six months
iv. Change in dietary intakes from baseline to six months
v. Self-reported perceptions of health from baseline to six months

#### b. Compliance Outcomes

i. Self-reported barriers to use of the powder collected at six months
ii. Adherence to the consumption regimen for the regular use of the powder
iii. Palatability (i.e. odour, colour, and taste assessments) collected at six months

#### d. Economic outcomes

i. Overall health status and health state values (using the EuroQol-5 Dimension EQ-5D) collected at six months for use in cost effectiveness analysis. Cost-effectiveness analysis will assess the effect of the intervention on health outcomes using quality-adjusted life years (QALY)

#### e. Safety outcomes

i. Adverse events (AEs)/harms monitoring for the duration of the trial and up to four weeks after last visit for unresolved AEs

### 3. Exploratory objectives

To explore the effects of six months of home beverage fortification with an iron and zinc enriched powder versus placebo in iron deficient reproductive aged women on:

a. The gut microbial metagenome at baseline, three months, and six months
b. The gut metabolome at baseline, three months, and six months

## Trial design [8]

This is a randomised, double-blind, placebo-controlled clinical trial conducted at a single centre in Ottawa, Canada. The trial aims to evaluate the efficacy of a mineral-enriched powder in biologically female participants with ID. Two parallel study groups will be randomly assigned in a 1:1 ratio to either the mineral-enriched powder group or the placebo group to assess the efficacy of the mineral-enriched powder compared to placebo. Each group will have 45 participants for a total of 90 participants, with a recruitment target of 150 participants to account for attrition. The study duration for each participant will be approximately seven months, including up to one month for screening followed by six months of daily use of the mineral-enriched powder or placebo. The study will include a total of one virtual screening visit (between days −28 to −1) and three in-person visits at baseline (V1, day 0), midline (three months [V2], day 90 ± 3), and endline (six months [V3], day 180 ± 3) of the study (Figure 1). Serum ferritin concentrations will be measured at baseline, midline, and endline. Other variables will be captured throughout the study as described below and in Figure 2.

**Figure 1.**
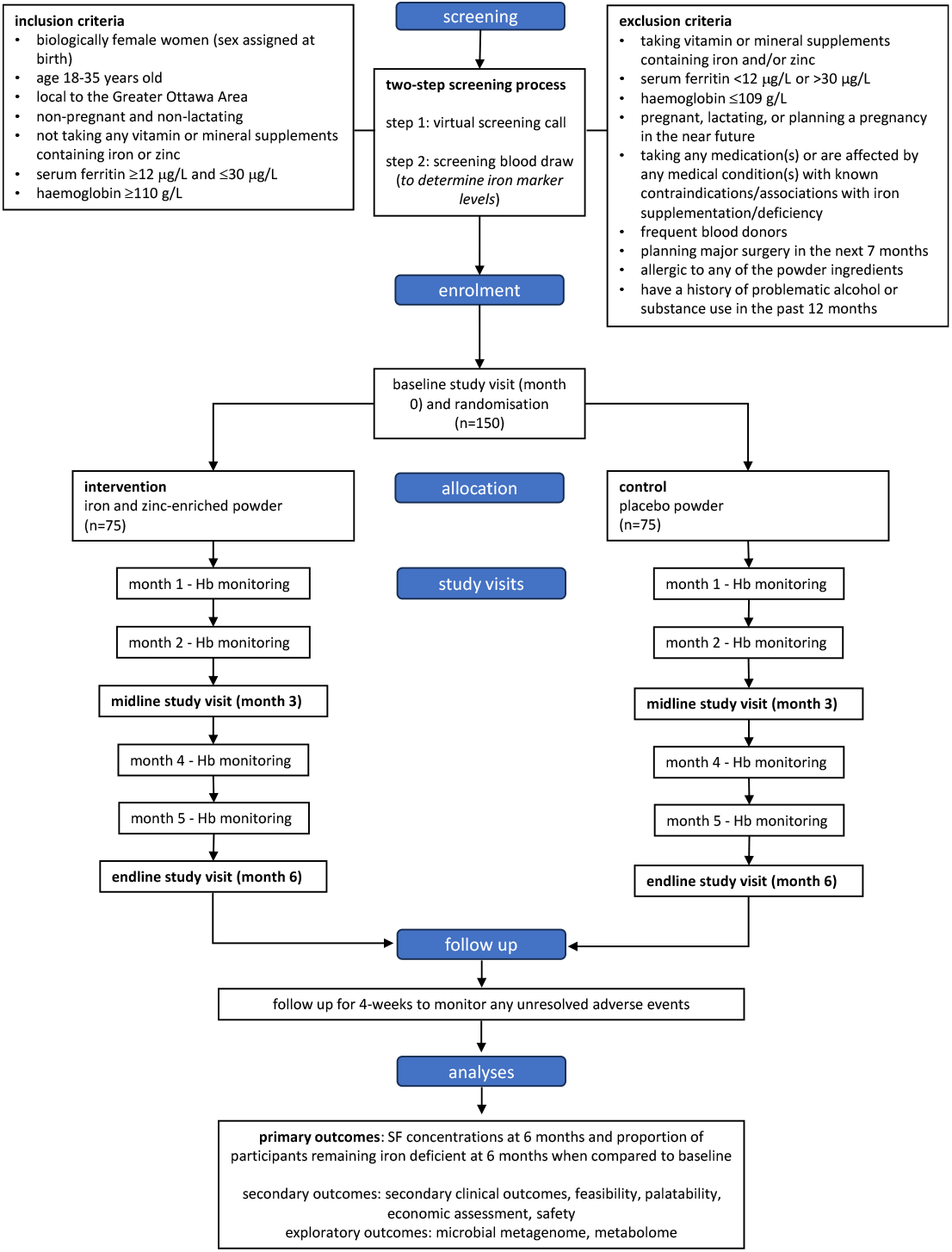
Participant flow diagram following the CONSORT-PRO^48^ guidelines.

**Figure 2.**
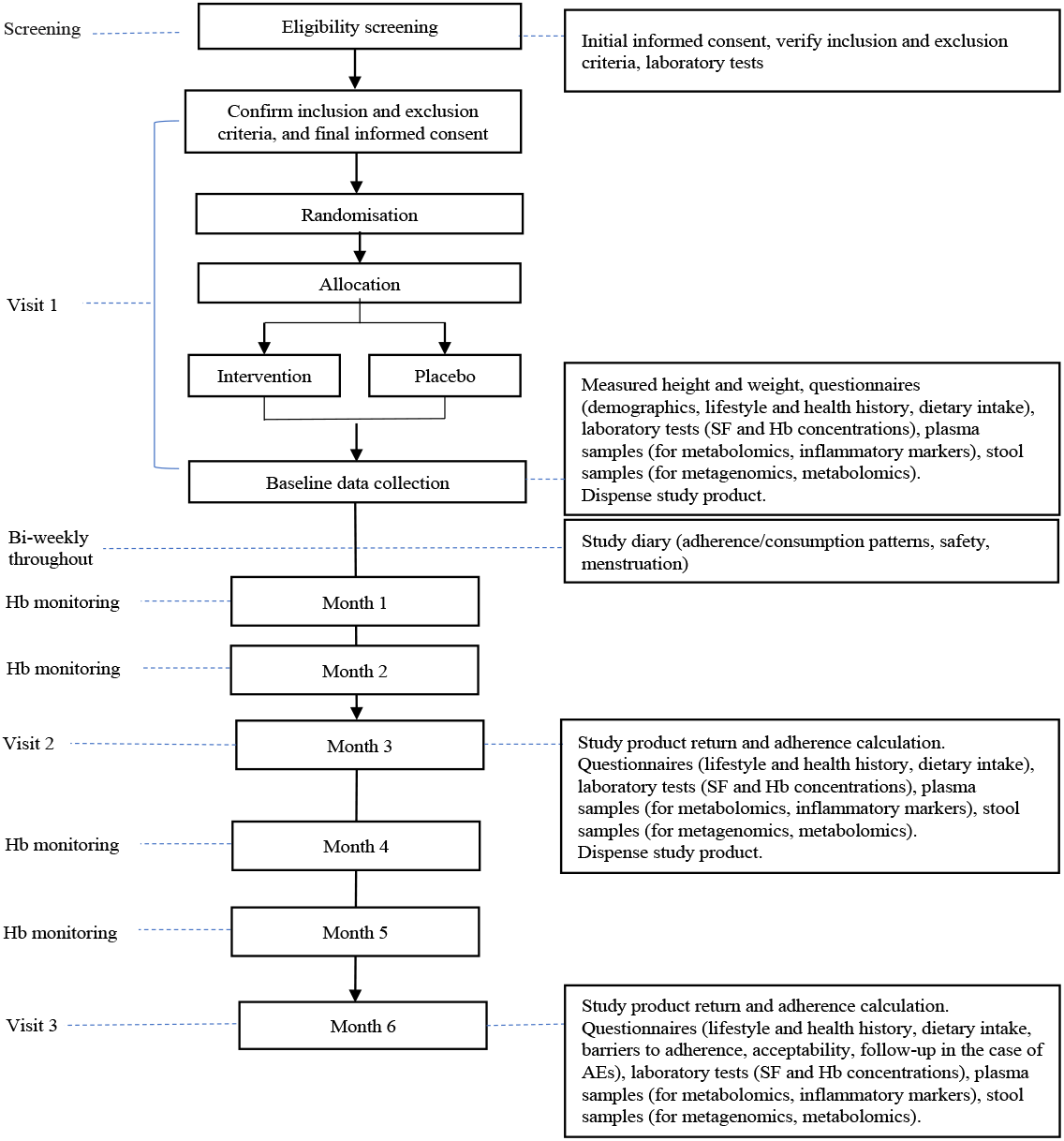
Schema of study timeline and assessments.

## Methods: Participants, interventions and outcomes

### Study setting [9]

Participants will be recruited from the greater Ottawa region in Ontario, Canada. All three study visits will be conducted at Carleton University. Blood samples for initial screening, primary outcome (i.e., SF concentrations), and secondary outcome (i.e., Hb, metabolomes and inflammatory biomarkers) measures will be collected at LifeLabs Medical Laboratory Services locations in the Ottawa area.

### Characteristics of the people who are needed for the trial

| Characteristic | The people we would expect to see included |
| --- | --- |
| Age | 18-35 years of age (inclusive) |
| Sex | Female. Females of reproductive age and pregnant people are at highest risk for developing ID and IDA due to their increased biological demand for iron to support menstruation and pregnancy. |
| Gender | Women; non-binary |
| Race, ethnicity and ancestry | The trial will aim for similar ethnic diversity as the recruitment area; given the convenience sampling approach, which includes advertisements at Carleton University and the University of Ottawa, may expect similar demographics as seen at these locations and surrounding areas. |
| Socioeconomic status | Recruitment in a high-income country, with universities as primary recruitment sites where various SES are likely to be represented in the student population. |
| Geographic location | Located in the greater Ottawa area and a resident of Ontario, Canada. |

### Eligibility criteria [10]

#### Inclusion criteria

- Non-pregnant, not planning to become pregnant during the study, and non-lactating
- English speaking with the ability to give informed consent
- Women who are biologically female
- 18-35 years of age (inclusive)
- Serum ferritin (SF) ≥12 μg/L and ≤30 μg/L
- Haemoglobin (Hb) ≥110 g/L
- Willing and able to agree to the requirements and lifestyle restrictions of this study
- Able to understand and read the questionnaires in English and carry out all study-related procedures
- Located in the greater Ottawa area and a resident of Ontario

#### Exclusion criteria

- Individuals who are not maintaining adequate birth control measures
- Have a known sensitivity, intolerability, or allergy to any of the study products or their excipients
- Are using vitamin and mineral supplements containing iron and/or zinc
- SF concentrations <12 μg/L or >30 μg/L
- Having moderate or severe anaemia (Hb ≤109 g/L)
- Expecting to change diet and exercise regimen in the next six months
- Are frequent blood donors
  - Have donated blood in the last four months
  - Donate blood more than two to three times per year
- Had major surgery in the past three months
- Have planned surgery during the course of the study
- History of problematic alcohol or substance use in the 12 months prior to screening (including having been hospitalized for such in an in-patient or out-patient intervention programme)
- Use of investigational product(s) in another research study within 30 days prior to the baseline visit or during the study duration
- Using drugs or medications with contraindications with iron supplementation
  - Antacids or proton pump inhibitors, H2 blockers
  - Salicylates, aspirin, corticosteroids, nonsteroidal anti-inflammatory drugs
  - Anticoagulants, antiplatelet compounds
  - Drugs with known contraindication with iron supplementation or fortification
  - Antiviral medications
  - Levothyroxine (Synthroid)
- Known medical history of specific conditions
  - Gastrointestinal disorders: celiac disease, ulcerative colitis, and Crohn’s disease
  - Gastric cancer and gastric polyps
  - Colon cancer
  - Diverticular bleeding
  - Inflammatory bowel diseases
  - Angiodysplasia
  - Helicobacter pylori infection
  - Hookworm (Ancylostoma duodenale and Necator americanus)
  - Defects of hemostasis (hereditary hemorrhagic telangectasia, von Willebrand disease)
  - Gastrectomy, duodenal bypass, bariatric surgery
  - Erythropoiesis-stimulating agent therapy
  - Chronic kidney disease
  - Hemochromatosis
  - Hemoglobinopathies
  - Blood clotting disorder
- Have any other active or unstable medical conditions or use of medications/supplements/therapies that according to the scientific literature, may adversely affect the participant’s ability to complete the study or its measures or pose a significant risk to the participant or the quality of the study data

### Who will take informed consent? [26a]

Informed consent will be obtained from participants by a research staff member. The consent process will occur in two stages (screening stage and enrolment stage). In the first stage, initial informed consent will be obtained from prospective participants both electronically and verbally. This consent will allow them to participate in the study screening process, which involves a questionnaire and a blood draw.

In the second stage, eligible and interested participants will be invited to attend their baseline study visit, in-person at Carleton University (Ottawa, Ontario). During the baseline study visit, participants will be debriefed on the study objectives, requirements and activities, and potential risks and benefits of participating in the study. Once the participant has been debriefed, they will be provided with a hardcopy of the final informed consent form to read and review before starting the study visit, assessments, and data collection. Once the consent form has been reviewed in its entirety, both the participant and research team member will sign and date two hardcopies of the final consent form, completing the informed consent process.

### Additional consent provisions for collection and use of participant data and biological specimens [26b]

During the informed consent process (second stage), participants will be informed that their anonymous (de-identified) data and biological samples will be kept by the research team to support future research studies on similar diseases or conditions. De-identified data and biological samples will be kept at Carleton University for 15 years after the study ends, in accordance with Health Canada regulations.

## Interventions

### Intervention description [11a]

The active intervention will be a powder containing maltodextrin, acetic acid (vinegar), sodium bicarbonate (baking soda), soluble ferrous iron (5 mg), and zinc sulfate monohydrate (10 mg). Participants will consume one sachet (1 g) of the study powder dissolved in 1L of water each day. All the ingredients within the formulation are approved for human consumption by Health Canada, either as food stabilisers or as food fortification (iron and zinc in the formulation).

### Explanation for the choice of comparators [6b]

The comparator will be a placebo powder containing maltodextrin, acetic acid (vinegar), and sodium bicarbonate (baking soda). The placebo powder will be identical to the active intervention powder in appearance but will not contain the active intervention ingredients (electrolytic iron and zinc sulfate monohydrate).

### Criteria for discontinuing or modifying allocated interventions [11b]

Stopping guidelines include pregnancy and serious adverse events (SAEs). Participants who become pregnant during the study will be instructed to stop taking the powder and will be withdrawn from the study immediately. Any reports of SAEs will be reported to the principal investigators (PIs) and the study physician throughout the study. The PIs and the study physician will determine if the participant should be removed from the study on an individual basis.

Participants will also be withdrawn and discontinue use of the powder due to worsening iron status (as defined by a decrease in Hb by > 5% from baseline concentrations, or SF/Hb below minimum inclusion levels [SF <12 μg/L and Hb ≤109 g/L]). Due to the fact that this study involves a blood draw for assessment of SF and Hb concentrations, there is the possibility of clinically significant or critical incidental findings. In the event of incidental clinical findings, participants may be withdrawn on recommendation of the study physician.

### Strategies to improve adherence to interventions [11c]

Acceptable levels of adherence will be determined by the following: individuals are consuming the product at least four days per week or a minimum of every other day for the duration of the trial. Adherence will be monitored in the bi-weekly study diary, where participants will describe their study product consumption patterns (frequency and amount). If a participant does not meet the acceptable level of adherence for one to two weeks, they will receive an adherence reminder from the research team. If the participant does not reach the acceptable level of adherence for more than two weeks, they will be withdrawn from the study. Participants will also be asked to return all used (opened) and unused (unopened) powder sachets at their three- and six-month study visits (visit 2 and visit 3). Adherence will also be measured based on the proportion of unused sachets.

### Relevant concomitant care permitted or prohibited during the trial [11d]

Participants will be asked to not consume any vitamin and mineral supplements containing iron and/or zinc for the duration of the study. Participants who choose to participate in the study will not receive standard of care treatment (e.g., supplemental iron) for their iron deficiency for the duration of the trial.

### Provisions for post-trial care [30]

Upon completion of the study, participants will be advised to contact their family physician or go to any walk-in clinic to follow up and monitor their iron status. Participants who are withdrawn from the study due to a decline in their iron status will also be referred to their family physician or qualified health care provider at a medical clinic. In the event that the participant cannot access their family physician or medical clinic in a timely manner, the study physician will recommend interim treatment options.

Participants who complete the study will be offered a personalised counselling session with a registered dietitian regarding dietary guidance on optimising their dietary intake of iron. Participants who are randomised to the placebo group will be contacted by the delegated unblinded staff and offered a 6-month supply of the active intervention product upon final completion of the study.

### Outcomes [12]

#### 1. Primary clinical outcomes

a. Serum ferritin (SF) concentrations at six months compared to baseline.
b. Proportion of participants remaining iron deficient at six months.

#### 2. Secondary outcomes

a. Clinical
  i. Change in SF and Hb concentrations from baseline, to three months, and six months.
  ii. Levels of metabolites (circulating metabolome) at baseline, three months, and six months.
  iii. Concentrations of key circulating pro-inflammatory biomarkers at baseline, three months, and six months.
  iv. Dietary recall: proportion of participants meeting dietary reference intakes, dietary patterns (e.g. vegetarian or vegan diets), Healthy Eating Index (HEI)/Health Eating Food Index (HEFI), and Dietary Inflammatory Index (DII) at baseline, three months, and six months.
  v. Self-reported perceptions of health at baseline and six months.
b. Feasibility
  i. Adherence to the consumption regimen for the regular use of the powder at three months and six months.
  ii. Self-reported barriers to use of the powder at six months.
c. Palatability
  i. Palatability (i.e. odour, colour, and taste assessments) at six months.
d. Economic
  i. Overall health status (EuroQol-5D visual analogue scale [EQ-5D VAS]), health state values (EuroQol-5D [EQ-5D]), quality-adjusted life years (QALYs), and cost-effectiveness analysis.
e. Safety
  i. Proportion of AEs and proportion of withdrawals from the study due to harms at baseline, three months, six months, and up to approximately seven months (four weeks post-study end).

#### 3. Exploratory outcomes

a. Experimental
  i. Gut microbial metagenome and exploratory analyses of inferred function of the metagenome at baseline, three months, and six months.
  ii. Gut metabolome at baseline, three months, and six months.

### Participant timeline [13]

### Sample size [14]

Group sizes of n=45/arm (total n=90) are required to detect a large effect size (d=0.6) in the difference in SF concentrations between the active intervention and control groups, assuming a power of 80% and a type 1 error probability of 5%^49^. This calculation is based on a t-test (two-sided) of the null hypothesis that there is no difference between the intervention and control groups. The number needed to recruit is 150 participants (75 intervention and 75 control), accounting for 40% loss/dropout. Similar studies have anticipated attrition ranges between 10-40%^38,50–53^, and we aimed to be conservative in estimated participant loss, given that iron levels are dynamic and may fluctuate between screening and baseline assessments. The number needed for data completion of primary outcome is 90 participants (45 intervention and 45 control).

### Recruitment [15]

Participants will be recruited using a convenience sampling approach through advertisements on social media, Carleton University, University of Ottawa, and Algonquin College campus, and Montfort Hospital. Study information will also be shared at health care clinics, centres such as gyms, local businesses, student housing, and community and promotional events. In addition, referrals from enrolled participants who are committed to the study will be encouraged. The research team members’ networks and word-of-mouth will also be used to reach out to prospective participants.

## Assignment of interventions: allocation

### Sequence generation [16a]

Participants will be randomised to the intervention and the placebo arms in a 1:1 allocation ratio using a computer-generated randomisation list using Research Electronic Data Capture (REDCap)^54,55^. The randomisation sequence was generated using block randomisation (block size of 10) with StataSE statistical software (version 16.1) and allowed for a 50% chance of random assignment to the intervention or the placebo group. Randomisation will take place following enrolment in the study, at the baseline study visit after the participant has completed the informed consent process.

### Concealment mechanism [16b]

A delegated unblinded staff will have access to the allocation codes for each arm. The allocation code will be concealed from the participants and the study personnel. Participants, study personnel, and data analysts will be unaware of arm type to which the participant has been randomised.

### Implementation [16c]

Participants will be randomised to groups ‘A’ or ‘B’ (randomisation codes) in REDCap following enrolment in the study, at the baseline study visit after the participant has signed the informed consent form.

## Assignment of interventions: Blinding

### Who will be blinded [17a]

Participants, study personnel, and data analysts will be blinded. Assignment of participants to intervention or placebo will be known only to delegated unblinded staff.

### Procedure for unblinding if needed [17b]

During the study, unblinding will not occur except in the case of an emergency, reportable SAE, or any other medical situation. Upon completion of the study, planned unblinding will occur following database lock when no further changes will be made to the dataset, definition of study populations, and a blinded statistical analysis.

## Data collection and management

### Plans for assessment and collection of outcomes [18a]

Data will be collected in three ways: First, biological samples and data collected during study visits (including anthropometric measures, adherence measures based on proportion of used and unused sachets, and blood measures) will be recorded in REDCap by the research team. Second, participant self-reported data following study visits (including sociodemographic data, lifestyle and health history factors, self-reported adherence, adverse events, menstrual patterns, and powder acceptability and palatability) will be collected using online study questionnaires sent directly to study participants via a password-protected research platform (REDCap) on a secure server hosted at the Montfort Hospital, Ottawa, ON. Third, dietary intake data will be collected using an online dietary recall software, the Automated Self-Administered 24-hour (ASA24), which measures all food and beverages consumed over a 24-hour period^56^. ASA24-CANADA 2018 is a Canadian version adapted to reflect the Canadian food supply, including portion sizes, and nutrient composition. These data will be exported as an anonymised Excel file.

### Plans to promote participant retention and complete follow-up [18b]

In order to increase participation, retention, and data completion rates, participants will be compensated in the following ways:

1. Participants who complete both screening steps of the screening stage (virtual screening and blood draw) will be entered into a draw for one of two $50 gift cards.
2. Participants who are enrolled in the study will be given a $50 gift card in compensation at the time of each in-person visit completion (e.g. completing questionnaires and providing biospecimens at the baseline, three-month, and six-month visits, for a total of $150). Additionally, their parking/travel fees for each visit will be reimbursed, up to $10/visit, with proof of receipt.
3. Enrolled participants will also be entered into a draw for one of two $100 grocery gift cards. Participants will earn a draw entry for each completed visit/procedure, for up to 7 total draw entries per participant (e.g. completing baseline, three-month, and six-month visit activities, and completing month 1, 2, 4, and 5 haemoglobin monitoring visits).
4. Participants will receive monthly newsletters containing dietary guidance developed by a registered dietitian on optimising dietary intake of iron.
5. Participants who are randomised to the placebo group will be contacted by the delegated unblinded staff and offered a six-month supply of the active intervention product upon final completion of the study.
6. Participants will be offered a counselling session with a registered dietitian, upon completion of the study. The consultation will include personalised dietary guidance based on their 24-hour recalls on increasing and optimising their dietary intake of iron.

### Data management [19]

#### Data management during the study

During the study, all study data will be collected or transmitted to and stored in REDCap electronic data capture tools hosted on the Montfort Hospital secure server^54,55^. Data collection will be the responsibility of the clinical trial staff. Electronic study source data collection forms will be created in REDCap. The data system includes password protection and internal quality checks, such as to identify data that appear incomplete. Clinical data will be entered directly from the source documents by the site staff.

Source Document Verification (SDV) will be completed by verifying the accuracy of data manually recorded in the electronic database against the original source document (electronic form; e.g., participant blood draw reports) by the research team, as detailed below. Any discrepancies noted between the source documents and data recorded in the electronic database will be addressed by the PIs. Any problems experienced by the site related to data entry or case report form completion will be reported to the sponsor’s project manager.

One hundred percent (100%) SDV for randomised participants will be performed on SF and Hb data.

Twenty-five percent (25%) SDV will be performed on the following data points:

- Signed initial and final consent forms
- Eligibility criteria
- Study ID assigned at screening visit (screening number)
- AEs and SAEs
- Questionnaires (e.g. *palatability questionnaire*)
- Protocol deviations (e.g. consumption patterns described in the *bi-weekly study diary*)
- Adherence and sachet tracking
- Sample collection, processing, storage, and shipping

Ten percent of participants will be randomly selected for 100% SDV of the study data.

#### Data management after study completion

Following study completion, de-identified data (only containing participants’ study ID) will be stored and used for secondary analyses after the identifying information in the database has been destroyed. Data will be stored in two places, including the electronic database on REDCap, our password-protected research database on the secure server hosted at the Montfort Hospital and accessible from a password protected computer. Data will also be exported in Excel or another research/statistical software to a password protected file, stored in a password-protected folder on the secure server hosted by Carleton University (and maintained by Research IT). The files will only be accessible to the principal investigators and research personnel responsible for analysing and interpreting the data.

Data will be retained and stored for the purpose of primary and secondary analyses and only individuals who are part of the research team and on the project ethics’ approval will be granted access to the stored data. The stored data will not be available to anyone for any purpose beyond the scope of the proposed trial.

In keeping with Health Canada regulations, electronic data, paper-based data (e.g., consent forms) and biological samples will be deleted after 15 years of completion of the study. Biological samples will be disposed of according to best practices for the disposal of biohazardous material at the time of the conclusion of the study. No records will be destroyed without the written consent of the sponsor.

### Confidentiality [27]

Participant confidentiality and privacy is strictly held in trust by the PIs, their staff, and the sponsor.

All participant data, including complete questionnaires, and their identifying information will be saved in REDCap. A unique study identification (study ID) will be generated for each participant to ensure anonymity. Once the trial is completed, only de-identified data will be exported from REDCap. The database will be exported in Excel or another research software to a password protected file, stored in a password-protected folder only accessible to selected research members (i.e. PIs, highly qualified personnel) on the secure server hosted by Carleton University.

Physical hardcopy documents will include signed final informed consent forms. Copies of physical documents, including informed consent forms, will be stored in locked filing cabinets in locked rooms at Carleton University. Copies of those documents will also be scanned and uploaded on REDCap.

Reports and presentations will include anonymous and aggregate data only.

### Plans for collection, laboratory evaluation and storage of biological specimens for genetic or molecular analysis in this trial/future use [33]

#### Stool samples

Stool samples will be self-collected by participants using a validated collection kit at baseline, three months, and six months. The samples will be delivered to Carleton University, either by the participant or using standard shipping procedures for biohazard samples. Stool samples will be used to assess the gut microbiome and microbial metabolome via standard procedures. Stool samples will be stored at −80°C at Carleton University until analysis.

#### Blood samples

Blood will be drawn by venipuncture at any LifeLabs Medical Laboratory Services location in the Greater Ottawa area at baseline, three months, and six months. Three vials of blood will be collected (for a combined volume of 13 mL; 4 mL for SF, 5 mL for Hb, and 4 mL for additional analyses). Approximately 2 mL of plasma (in 400 µL aliquots) will be sent to the research team at Carleton University for analysis of the circulating metabolome (via metabolomics approaches) and inflammatory biomarkers (via protein/immunological assays). The samples will be frozen and delivered to Carleton University using standard shipping procedures for biohazard samples, in accordance with LifeLabs’ shipping guidelines. Plasma samples will be stored at −80°C at Carleton University until analysis.

On the months that participants are not having full laboratory blood draws to determine SF and Hb concentrations at LifeLabs Medical Laboratory Services (months 1, 2, 4, and 5), a capillary blood sample will be collected via pin-prick blood collection for safety monitoring of Hb concentrations. Capillary blood samples will be collected by a member of the study team at Carleton University or at a location that is most convenient for the participant. Samples will be collected according to the WHO’s guidance document and procedure for collecting capillary blood samples^57^.

## Statistical methods

### Statistical methods for primary and secondary outcomes [20a]

Participant baseline demographics will be reported descriptively for intervention and placebo groups. The intervention and placebo groups will be compared for primary and secondary outcomes, reporting continuous variables as mean difference (95% CI), means and standard deviations (SD) or medians and interquartile ranges, and categorical variables as proportion (with counts and percentage). Primary outcomes are intended to establish effectiveness of the intervention. Secondary outcomes are intended to support primary outcomes and/or demonstrate additional effects.

Both intention-to-treat (ITT) and per-protocol (PP) population analyses will be performed. The ITT population will include all randomised participants who received at least one dose of study product as per the International Council for Harmonisation Good Clinical Practice (ICH GCP) guidelines^58,59^. The PP population will include all randomised participants who received at least one dose of the study product *and* adhered to the study protocol for the duration of the study (e.g., adherence to study product, provide primary outcome data [SF concentrations at baseline and 6 months]).

#### Assessment and analysis of primary outcome

The primary endpoints are defined as serum ferritin concentrations at six months and the proportion of participants in the intervention and placebo group who are no longer iron deficient after six months. Serum ferritin levels will be adjusted for the effects of inflammation (C-reactive protein and α-1-acid glycoprotein), using the Biomarkers Reflecting Inflammation and Nutritional Determinants of Anemia (BRINDA) method^60,61^, as SF levels can be transiently elevated in response to inflammation, potentially resulting in underestimates of ID. Difference in mean SF concentrations between placebo and active intervention groups at six months will be determined using the independent samples t-test (or Welch or Mann-Whitney U tests for unequal variance or nonnormality). Difference in proportion of participants with low SF (remain ID) between groups at six months will be determined using the chi-square test. We will also report risk of low SF (ID) at six months using risk ratio (RR). Change in proportion with low SF between baseline and six months within each group will be determined using the McNemar test.

#### Assessment and analysis of secondary outcomes

The secondary endpoints are categorised as clinical, feasibility, palatability, economic, and safety outcomes.

#### Clinical: iron status, circulating metabolome, plasma inflammatory markers (pro-inflammatory cytokines), nutrition, and self-reported perceptions of health

Active intervention and placebo groups will be compared for iron status (SF and Hb levels), levels of circulating metabolites, and levels of pro-inflammatory cytokines at baseline, three- and six-month timepoints.

Data will be analysed using both unadjusted (univariate) and adjusted (multivariate) linear regression models, and results from both models will be reported. Adjusted models will consider potential covariables including baseline SF concentrations; iron intake; menstrual patterns; and physical activity variables. Change in SF and Hb concentrations from baseline, three months, to six months will be analysed using longitudinal analysis of covariance to estimate the treatment effect, controlling for baseline values.

#### Dietary intakes

Active intervention and placebo groups will be compared for mean dietary iron intake (obtained from the ASA24 for Canada) at baseline, three, and six months timepoints using the independent samples t-test (or Welch or Mann-Whitney U tests for unequal variance or nonnormality). Difference in proportion of participants having met estimated average requirements (EAR) between intervention and placebo groups will be determined using the chi-square test. Active intervention and placebo groups will be compared for mean HEI/HEFI and DII at baseline, three-, and six-month timepoints. Dietary intake estimates will be corrected for measurement error inherent to 24-hour recall data using the National Cancer Institute (NCI) method. We will also assess the effect of dietary pattern (e.g. vegan/vegetarian/omnivore) on iron intake and status. Data will be analysed using both unadjusted (simple) and adjusted (multivariable) linear regression models, and results from both models will be reported. Only participants with complete dietary intake data (a minimum of 2 days of 24-hour recall per timepoint) will be used for analysis.

#### Perceived health

Difference in proportion of participants reporting improved perceived health (obtained from the lifestyle and health history questionnaire) between active intervention and placebo groups at six months will be determined using the chi-square test. Change in proportion reporting improved perceived health between baseline and six months within each group will be determined using the McNemar test.

#### Barriers to use

Active intervention and placebo groups will be compared for barriers to the use of the powder (data obtained from the barriers to adherence questionnaire) at six months. Difference in proportion of participants reporting perceived barriers between intervention and placebo groups will be determined using the chi-square test. We will report a qualitative summary of the different types of perceived barriers indicated and associations between proportion of participants reporting perceived barriers with adherence to the study regimen and overall health status using chi-square test and contingency analysis.

#### Adherence to use

Active intervention and placebo groups will be compared for adherence (data obtained from the bi-weekly study diary) to consumption regimen at three and six months. Difference in proportion of participants reporting >80% adherence (defined as preparation and consumption of the intervention powder on at least 4 days per week or a minimum of every second day throughout the trial), and proportion of unused sachets, between intervention and placebo groups will be determined using the chi-square test. We will report associations between proportion of participants reporting >80% adherence with SF levels, proportion of participants who remain ID, and overall perceptions of health outcomes.

#### Palatability

Active intervention and placebo groups will be compared for palatability assessments of the powder solution and its overall acceptability at six months (obtained from the palatability and acceptability questionnaire) using a 9-point hedonic scale (where ≥5 indicates that the solution was liked). Differences in participant palatability scores (for colour, odour, sweet, sour, bitter, salty, umami and overall liking) between intervention and placebo groups will be determined using Wilcoxon-Rank Sum test (or Welch or Mann-Whitney U tests for unequal variance or nonnormality) and visualised using polar plots. Proportion of participants who indicate they would be willing to drink the sample daily will be determined using the chi-square test.

#### Economic

Active intervention and placebo groups will be compared for overall health and health improvements at six months using the EQ-5D VAS. Mean index values will be used to calculate quality-adjusted life years (QALYs) and perform cost-effectiveness analysis. The EQ-5D VAS score represents the patient perspective about the value of the health state while the Index values represent the societal perspective (what the general population thinks about the value of the health state). Differences in mean EQ-5D VAS score and QALYs between intervention and placebo groups at six months will be determined using independent samples t-test (or Welch or Mann-Whitney U tests for unequal variance or nonnormality).

#### Safety

We will compare active intervention and placebo groups for adverse effects (AEs) and withdrawals due to harms descriptively, reporting proportion (with counts and percentage) and ratio/relative risk at three and six months. AEs will be visualised using forest plots and time-to-event plots.

Safety data will be collected from participants who received at least one day of the active intervention or placebo from baseline to six months and up to four weeks after the last study visit for unresolved AEs (data obtained from the bi-weekly study diary). Difference in proportion of participants reporting AEs and in proportion of withdrawals due to harms between intervention and placebo groups will be compared at three and six months, and at four weeks after the last study visit using chi-square tests. We will also report risk of AEs at three and six months, and four weeks after the last study visit, using risk ratio (RR). We will report data relevant to AEs including descriptions, severity, duration, timing, and demographic/health information of the reporting participant.

#### Assessment and analysis of exploratory outcomes

The exploratory outcomes will investigate the effect of the intervention on the gut microbiome (data obtained from stool samples) by exploring the effects on the microbial metagenome and microbial metabolome. The gut microbiome is an emerging modifier of iron absorption and homeostasis^62,63^, and is known to respond to exogenous factors including diet and pharmacological agents^64,65^. Assessing the gut metagenome and metabolome in this trial will therefore provide critical insight into whether the iron-fortified intervention alters microbial composition, and whether microbiome variability contributes to individual differences in iron status and treatment response.

#### Microbial metagenome

We will compare placebo and intervention groups for experimental (microbiome) outcomes descriptively, reporting relative taxonomic abundance, diversity and variance following standardised pipelines^66,67^. Exploratory analyses of inferred function of the metagenome will be undertaken using compositional approaches^68,69^ and visualised using compositional and scale-aware approaches to minimise false discovery rates^67,70,71^. Differentially abundant microbial functions and pathways between active and placebo groups will be determined (ALDEx2 and/or ALDEx3^72^) and network/pathway analysis will be conducted using Ingenuity Pathway Analysis (IPA) and HUMAnN3. This will generate data on genes, networks and biological pathways/processes that may be influenced by the intervention, accounting for clinical data.

#### Microbial metabolome

We will further assess microbial function^73^ (and relate to computationally-derived functions from metagenome data) by measuring functional metabolites (microbial products, short chain fatty acids, vitamins, metals/minerals). Data will be processed under standardised pipelines with visualisation and pathway analysis by Agilent Mass Profiler (using AMP Professional Software), MetaboAnalyst, KEGG database, GeneOntology (GO – analysed using ALDEx2 and/or ALDEx3)^72,74,75^. Differentially abundant candidate metabolites/metabolic pathways between active and placebo groups will be determined (including using IPA), and untargeted analysis of metabolites/metabolic pathways will be conducted (e.g. using the MetaboDynamics R package^76^).

#### Reporting of outcomes

We will compare intervention and placebo groups for outcomes, reporting continuous variables as means and standard deviations (SD) or medians and interquartile ranges, and categorical variables as proportion (with counts and percentage). Primary outcomes are intended to establish effectiveness of the intervention. Secondary outcomes are intended to support primary outcomes and/or demonstrate additional effects. Relationships between outcomes will be reported (in supplementary tables/appendices) to provide information that could inform outcome selection in future research and/or provide insight into possible mechanisms, measured variables and constructs^77^. Table 1 summarises the outcomes, measurements and analyses.

**Table 1.**
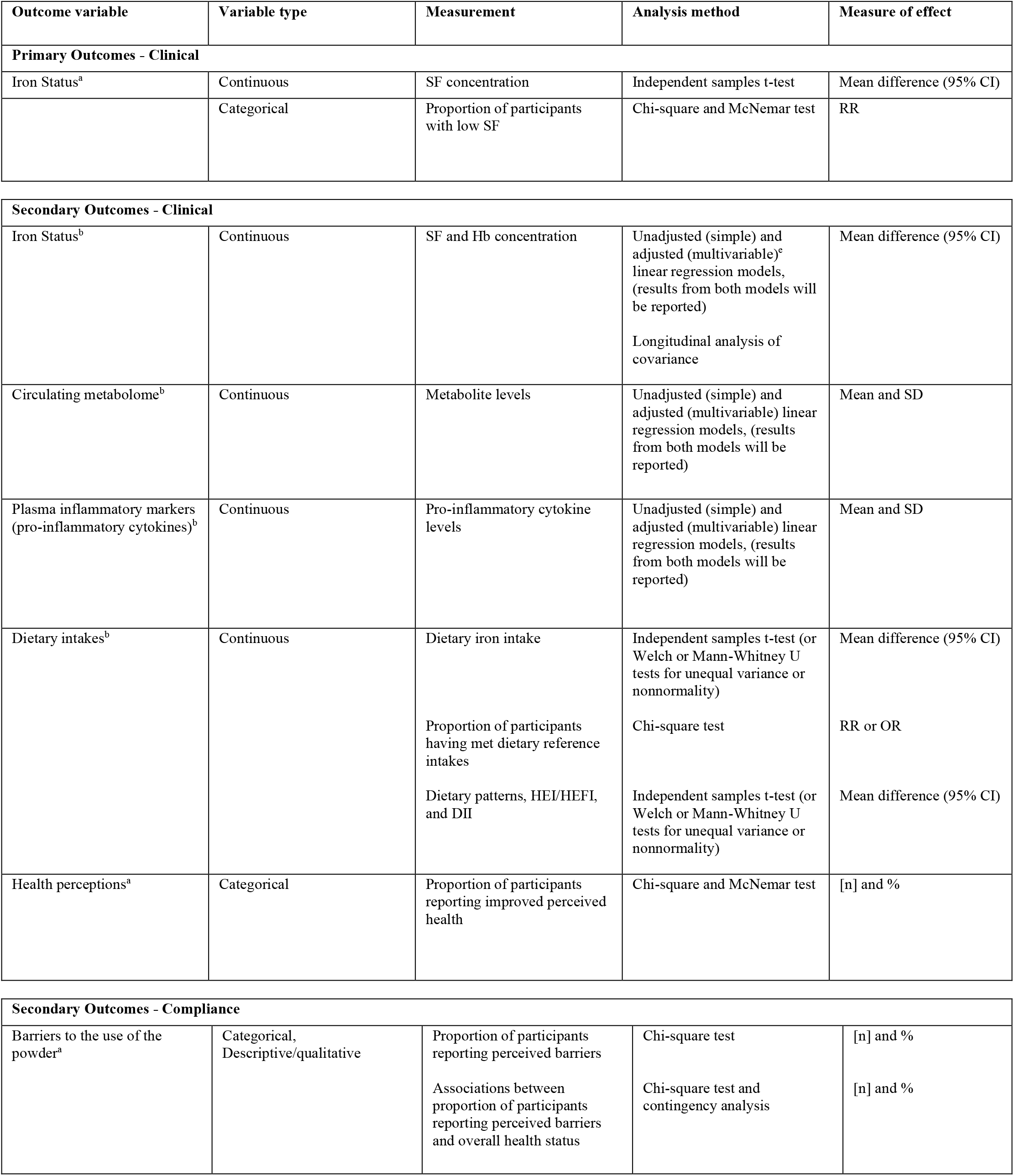
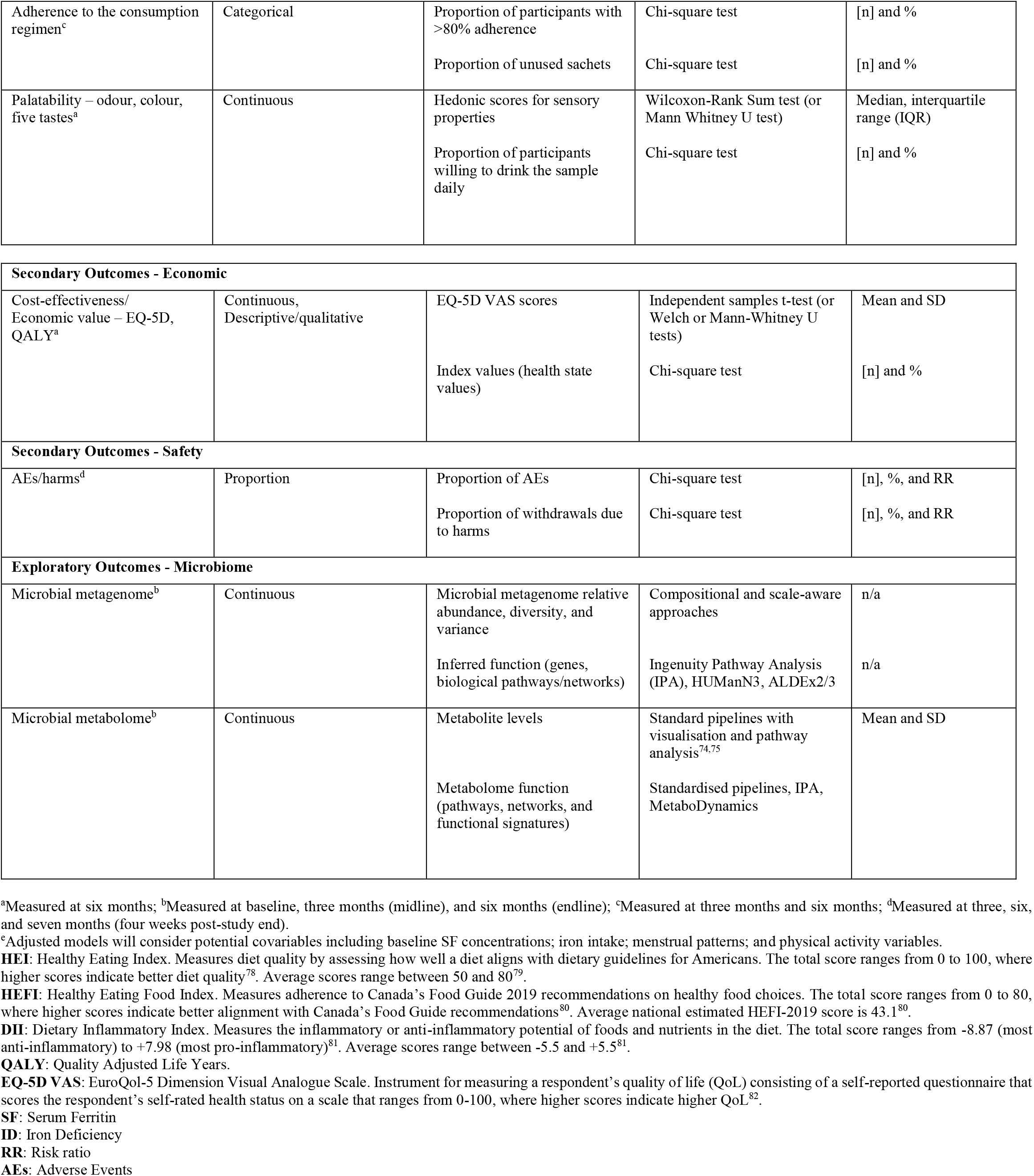
Summary of outcomes, measurements and analyses.

### Interim analyses [21b]

Interim analyses will be conducted on the primary outcome data to check for data validity, accuracy, and potential outliers. Interim analyses may also be conducted for safety purposes.

### Methods for additional analyses (e.g. subgroup analyses) [20b]

For participants who have *screening* serum ferritin levels within the inclusion range (SF ≥12 µg/L and ≤30 µg/L; considered ID), but at *baseline*, have SF levels >30 ug/L, a sensitivity analysis will be performed (for participants with baseline serum ferritin levels ≤30 µg/L only, and inclusive of participants with baseline serum ferritin levels >30 µg/L).

### Methods in analysis to handle protocol non-adherence and any statistical methods to handle missing data [20c]

If 5% or more of the total data are missing, multiple imputation (MI) approaches will be used to account for the primary and secondary outcomes if the data is missing at random (MAR) using a statistical model that takes into account observed data and other relevant variables^83–85^. By using multiple imputation approaches, we aim to obtain unbiased estimates of treatment effects and minimize potential bias.

### Plans to give access to the full protocol, participant level-data and statistical code [31c]

This trial protocol is registered at clinicaltrials.gov (NCT0599016). Participant-level data will be retained and stored for the purpose of primary and secondary analyses and only individuals who are part of the research team will be granted access to the stored data. The stored data will not be available to anyone for any purpose beyond the scope of the proposed trial.

The statistical codes for analyses will be available on an open repository e.g. GitHub, Zenodo.

## Oversight and monitoring

### Composition of the coordinating centre and trial steering committee [5d]

Dr. Kristin Connor, PhD

*Principal Investigator*

Department of Health Sciences

Carleton University

Phone: 613-520-2600 ex. 4202

Dr. Bénédicte Fontaine-Bisson, PhD, RD

*Principal Investigator*

Faculty of Health Sciences

School of Nutrition Sciences

University of Ottawa

Phone: 613-562-5800 ex. 3482

Dr. Brianne Clouthier, MD, CCFP

*Qualified physician, responsible for all trial-site related medical decisions*

Renfrew Medical Group

Dr. Lawrence Mbuagbaw, MD, MPH, PhD

Department of Health Research Methods, Evidence, and Impact, McMaster University

Dr. Jessie Burns, PhD

*Research team member*

Department of Health Sciences, Carleton University

Hailey Scott, MSc

*Research team member*

Department of Health Sciences, Carleton University

Alexandra Rooney, RD

*Research team member and registered dietitian*

Department of Health Sciences, Carleton University

Clara Miller, BSc

*Research team member*

Department of Health Sciences, Carleton University

Hauna Sheyholislami, MSc

*Research team member*

Department of Health Sciences, Carleton University

### Composition of the data monitoring committee, its role and reporting structure [21a]

The Data Monitoring Committee (DMC) will be responsible for monitoring efficacy, clinical data, and conducting safety assessments. It will be comprised of the PIs, the study physician, the study research coordinator and the secondary data entry personnel. The DMC will address safety concerns as they arise based on reported clinical outcomes and safety data captured in the bi-weekly study diary. The PIs, study research coordinator, and secondary data entry personnel will meet weekly to review data on product safety, side effects, and clinical data from blood draws. The study physician will oversee blood results, and any SAEs will also be discussed with the study physician. The DMC may ask the designated unblinded staff to unblind data for the purpose of making safety recommendations based on interim analyses and will be responsible for determining if the study design should be modified, if a participant should be withdrawn from the study, or if the study should be terminated. The DMC will also be responsible for reporting any SAEs to the REB.

### Adverse event reporting and harms [22]

All AEs and SAEs will be captured in the bi-weekly study diary from the start of administration of the study product (day 0) until the last study visit (day 180 ± 3). The study diary will be reviewed by the research team and any SAEs will be reviewed by the study physician to determine if it is a serious adverse effect (if causality can be established). Participants will be instructed to report any AEs and SAEs in their bi-weekly study diary and to report SAEs to a member of the study team immediately. Participants will be instructed to discontinue the study product immediately in the event of a SAEs. Unresolved AEs or SAEs will be followed for four weeks after the participant has completed the study. Adverse effects (AEs for which a causal relationship between the intervention and the event is at least a reasonable possibility) will be reported to the sponsor using the Case Report Form (CRF). SAEs will be recorded and reported by the Investigator site to the Sponsor no later than 24 hours from the time the site became aware of it. The Principal Investigators will be responsible for reporting the SAE to the REB within 24 hours of the receipt of the initial and follow-up reports on SAEs. The Sponsor will be required to include any SAEs in their standard post-marketing SAE reporting process with Health Canada.

Reporting of AEs in publications will compare active intervention and placebo groups for AEs and withdrawals due to harms descriptively, reporting proportion (with counts and percentage) and ratio/relative risk. Data relevant to AEs including descriptions, severity, duration, timing, and any relevant demographic/health information of the reporting participant may also be reported.

### Frequency and plans for auditing trial conduct [23]

Quality Assurance audits may be performed on this study at the discretion of the Sponsor.

### Plans for communicating important protocol amendments to relevant parties (e.g. trial participants, ethical committees) [25]

Any amendment to the protocol (and, if applicable, the consent form) will require review and authorization/approval by the REBs before the changes are implemented to the study.

## Dissemination plans [31a]

Study results will be disseminated following CONSORT guidelines for randomised controlled trials^52^, including the extension for harms^86^, and extension for CONSORT-PRO^48^, with a participant flow diagram. Results will be published in scientific journals, shared at scientific conferences, and communicated to other knowledge users (health care professionals, the public, public health and policy stakeholders) through various knowledge translation and exchange activities and platforms. These may include the study website, research and educational outreach talks, and communications briefs. The sponsor places no restrictions on data dissemination.

## Discussion

Not applicable, no other practical or operational issues involved in performing the study to disclose.

## Data Availability

Not applicable. This manuscript does not report data generation or analysis.

## Trial status

Study start: April, 2023 (actual)

Primary recruitment completion: September, 2025 (actual)

## Abbreviations

Abbreviations and definitions

### Abbreviation Definition

AE: Adverse event
CONSORT: Consolidated Standards of Reporting Trials
CRF: Case report form
DMC: Data Monitoring Committee
EQ-5D: EuroQol-5 Dimension
Hb: Haemoglobin
ICF: Informed consent form
ID: Iron deficiency
IDA: Iron deficiency anaemia
KEGG: Kyoto Encyclopedia of Genes and Genomes
LIL: Lucky Iron Life
NGOs: Non-governmental organizations

### Abbreviation Definition

NNHPD: Natural and Non-Prescription Health Products Directorate
QALY: Quality-adjusted life year
REB: Research ethics board
REDCap: Research Electronic Data Capture
SAE: Serious adverse event
SD: Standard deviation
SDV: Source document verification
SF: Serum ferritin
SPIRIT: Standard Protocol Items Recommendations for Interventional Trials
Study ID: Study identification number
V: Visit

## Declarations

## Authors’ contributions [31b]

JLB: contributed to the design of the protocol, contributed to the writing of the manuscript, approved the final version of the manuscript to be published, and agrees to be accountable for the work.

HS: contributed to the review of the manuscript, approved the final version of the manuscript to be published, and agrees to be accountable for the work.

LM: contributed to the design of the protocol and plan for statistical analysis, contributed to the review of the manuscript, approved the final version of the manuscript to be published, and agrees to be accountable for the work.

CHM: contributed to the writing of the manuscript, approved the final version of the manuscript to be published, and agrees to be accountable for the work.

HS: contributed to the design of the protocol, contributed to the review of the manuscript, approved the final version of the manuscript to be published, and agrees to be accountable for the work.

BAC: contributed to the design of the protocol, contributed to the review of the manuscript, approved the final version of the manuscript to be published, and agrees to be accountable for the work.

BFB: designed the protocol, oversight of the operationalization of the study, contributed to the writing of the manuscript, approved the final version of the manuscript to be published, and agrees to be accountable for the work.

KLC: designed the protocol, oversight of the operationalization of the study, contributed to the writing of the manuscript, approved the final version of the manuscript to be published, and agrees to be accountable for the work.

## Availability of data and materials [29]

Not applicable. This manuscript does not report data generation or analysis.

## Ethics approval and consent to participate [24]

The study protocol was approved by the Montfort Hospital Research Ethics Board (#22-23-07-017), the Carleton University Research Ethics Board (#118907), the University of Ottawa Research Ethics Board (#H-01-23-8256), and the Algonquin College Research Ethics Board (#2024-Feb-Connor).

The study protocol has also received Clinical Trial Application Authorisation from Health Canada’s Natural and Nonprescriptive Health Products Directorate (protocol number: LIFe2022RCT2, file number: 257665).

## Consent for publication [32]}

Not applicable

## Authors’ information (optional)

Not applicable

